# Discordance in pleural mesothelioma response classification and modelling of impact on clinical trials

**DOI:** 10.64898/2026.03.18.26348731

**Authors:** Gordon W Cowell, Joshua Roche, Colin Noble, David B Stobo, Andrew Papanastasiou, Andrew C Kidd, Selina Tsim, Kevin G Blyth

## Abstract

**Introduction:** Agreement between radiologists regarding treatment response in Pleural Mesothelioma (PM) is acknowledged to be poor, but downstream effects in clinical trials have not been quantified.

**Methods:** We performed a mixed methods study, composed of a multicentre, retrospective cohort study and *in silico* modelling. CT images and data were retrieved from 4 UK centres regarding chemotherapy-treated patients. Expert radiologists classified response using modified Response Evaluation Criteria In Solid Tumours criteria (mRECIST) v1.1, generating discordance rate (%) and agreement. *In silico* modelling simulated two-arm trials of an active therapy with intended 80% power and confidence intervals for four endpoints (objective response rate (ORR), disease control rate (DCR), progression-free survival (PFS), overall survival (OS)) covering 95% of the true effect. Actual power and endpoint coverage were modelled against mRECIST misclassification rate (a single reporter equivalent of discordance rate). Consecutive simulations varied misclassification rate from 0-100% in 1% increments, each repeated 10,000 times.

**Results:** 172 cases were included. Discordance rate was 35% (60/172), kappa=0.456. *In silico* modelling demonstrated reduced power and endpoint precision with increasing misclassification. At 17% misclassification, corresponding to the observed 35% discordance, power dropped from 80% to 55% for ORR, 53% for DCR, 65% for PFS and 66% for OS, with endpoint coverage reduced to 88%, 89%, 92% and 92%, respectively. 50/60 (83%) discordances reflected interpretation or measurement differences intrinsic to mRECIST. Discordance was not associated with tumour volume.

**Conclusions:** Inconsistent response classification is common in PM and substantially reduces statistical power and endpoint precision in clinical trials.

## INTRODUCTION

Pleural mesothelioma (PM) is an aggressive thoracic malignancy associated with prior asbestos exposure. The diagnosis is often delayed due to subtle imaging features, which may be limited to minimal pleural thickening and a unilateral effusion in early-stage disease. In advanced disease, first-line systemic therapies, including platinum-pemetrexed chemotherapy^1^ and ipilimumab-nivolumab immunotherapy^2^ extend survival in a minority of patients, making clinical trials a high priority.

Efficient trial delivery relies on reliable radiological response assessment. In PM, this currently involves use of a modified version of the Response Evaluation Criteria In Solid Tumours criteria (called Modified RECIST v1.1)^3^ . This schema requires the reporting radiologist to make six unidimensional pleural tumour measurements, add these to measurements from any extra-pleural sites, and replicate the position of these measurements on follow-up scans. Unsurprisingly, previous studies demonstrate significant inter-observer differences using this method, which exceeded 30% in a study reported by Armato et al^4^ . Since the mRECIST thresholds for progression and response are ≥20% and ≤30% respectively, this variation could potentially result in discordant classifications by two readers based on the same CT images.

To our knowledge, the actual frequency of discordant classification has not previously been reported. These data are important for clinical trial design and in routine clinical practice, where multiple reporters are almost never used. In routine care, a single inaccurate response assessment may therefore lead to continuation of a toxic and ineffective drug or premature withdrawal of one that is having an effect. In this study, we set out to systematically examine the extent and sources of discordant response classification using mRECIST v1.1 in PM. We also modelled the impact of misclassified response on the actual statistical power and endpoint precision in clinical trials using four typical endpoints (objective response rate (ORR), disease control rate (DCR), progression-free survival (PFS), overall survival (OS)).

## METHODS

### Study Design & Objectives

We performed a mixed methods study, composed of a multicentre retrospective cohort study and *in silico* modelling. The primary objective was to determine the frequency of discordant response classification, defined as discordance rate (%) between two experienced thoracic radiologists using mRECIST v1.1^3^ . Our first secondary objective was to investigate, using *in silico* modelling, the impact of response misclassification on the statistical power and the precision of four typical trial endpoints: objective response rate (ORR), disease control rate (DCR), progression-free survival (PFS) and overall survival (OS). Additional secondary objectives were (a) to perform a root cause analysis of any discordance observed using the following themes: system errors, human errors, variances related to image interpretation and (b) to compare discordance rate with volumetric change in cases with this data available.

### Ethical Approval and Funding

Ethical approval was granted by the West of Scotland Safe Haven (Ref GSH/18/ON/001) as part of the PRiSM study, which set out to define and validate a genomic classifier of chemoresistance based on radiological response^5^ . The study was funded by British Lung Foundation (now Asthma + Lung UK), Grant Ref MPG16-7.

### Eligibility Criteria

Cases were eligible if they had received chemotherapy (cisplatin/carboplatin-pemetrexed) for PM at a participating centre (Glasgow, Wythenshawe or Leicester) and had both baseline (pre-chemotherapy) and response CT images available. The response CT had to be at least 4 weeks after chemotherapy initiation.

### Data Collection and CT Image Acquisition

Patient demographics, histological subtype, diagnosis date, chemotherapy regime and CT parameters, including contrast timing and scanner manufacturer, were extracted from electronic records. CT examinations were undertaken during routine care, on a variety of CT scanners. Although local imaging protocols varied, imaging was generally acquired in the portal venous phase (∼65 seconds after injection of 75–95 mL of iodinated contrast material).

### mRECIST Classification

CT images, in DICOM format, were pseudo-anonymised and transferred securely to Glasgow for reporting. Each CT pair (baseline and response) was reported by two expert reviewers (Reviewer 1: GWC and Reviewer 2: CN) who were blinded to other data. Both expert reviewers were members of the Scottish National Mesothelioma MDT, with 13 and 16 years of thoracic radiology experience, respectively. CT images were loaded onto the national Picture Archive and Communication System (PACS, Carestream Vue v12.2, Rochester, USA) for review and reporting using mRECIST 1.1. As previously described^3^ , this involved 6 unidimensional measurements of pleural tumour per scan. Measurements were made perpendicular to the chest wall at 3 different axial levels at least 1 cm apart. Measurement of any pathological nodes or extra-pleural sites of disease were also recorded. All measurements were entered into a MS Access® database. Tumour measurements were summed and each case classified into partial response (PR), progressive disease (PD) and stable disease (SD) groups. PR was defined as ≥30% reduction in total tumour measurements. PD was defined as ≥20% increase in total tumour measurements or the appearance of new sites of disease. SD was any outcome not meeting PR or PD criteria.

### Primary Analysis regarding Discordance Rate

Discordance rate was reported as a simple proportion (%). In discordant cases, a third experienced thoracic radiologist (DBS) provided an adjudicating opinion. In subsequent statistical analyses, Reviewer 1 measurements were used, except in the discordant cases where the correct response was defined by the classification with which Reviewer 3 agreed.

### Secondary Analyses

#### In Silico Modelling

An *in silico* simulation study was performed to assess how response misclassification affects observed power and the precision of four commonly used trial endpoints: objective response rate (ORR), disease control rate (DCR), progression-free survival (PFS) and overall survival (OS). This involved a series of simulated two-arm trials of an active PM therapy targeting either (a) an absolute difference in response rate of 20% for ORR and DCR models, or (b) an effective HR of 0.7 for models regarding PFS and OS. Each trial was designed to deliver 80% power using a threshold of statistical significance of p<0.05 and a confidence interval (CI) around the chosen endpoint that covered 95% of the true treatment effect. For OS models, we assumed that the effective hazard ratio (HR) was consistent with the intended power, without attenuation related to post-protocol therapies which can be an issue in the real world. Each iteration of the simulation represented a discrete clinical trial where the only feature altered was the misclassification rate, which was varied from 0 to 100% in increments of 1%, distributed randomly across the trial population. Individual patient survival times were modelled stochastically. Each simulation was repeated 10,000 times per misclassification rate, generating robust inferences and confidence intervals for the power and coverage at each misclassification rate. Patients in the simulated trials were followed up for 24 months for vital status, after which they were censored. For PFS models, protocolised imaging required continued for up to 24 months or progression, whichever occurred first.

Separate models and therefore separate plots were generated for ORR, DCR, PFS and OS. For models regarding PFS and OS, where response misclassification must be integrated with modelling of a clinical decision to stop or continue therapy based on observed response, five assumptions were made: (1) that there was no treatment interaction nor any prolonged treatment effect after stopping the trial drug, at which point patients switched to symptom directed care; this was deemed appropriate given the limited second-line options in PM^6^ (2) the median OS following cessation of the trial drug was 8 months; this was extrapolated from the median time from progression to death in EMPACIS (platinum-pemetrexed chemotherapy: 6 months^1^ ) and CM743 (ipilimumab-nivolumab immunotherapy: 11 months^2^) (3) the probability of misclassification is the same for each of the mRECIST classes (PR, SD, PD), i.e. a non-differential misclassification (4) median PFS on standard of care was 5.6 months, based on the symptom directed care arm plus chemotherapy in the MS01 trial ^7^ (5) median OS when remaining on an ineffective treatment was 3.4 months, based on Blayney et al, in which the median OS was 3.4 months in patients who experienced PD on receipt of first-line chemotherapy^8^ . The illness-death structure along with the PM-specific parameters produce an individual-level Kendall’s tau of approximately 0.50, which is consistent with the tau of 0.45–0.47 reported by Wang et al. in their analysis of 716 patients across 17 phase II trials^9^ . This alignment provides reassurance that the modelling process inherently reflects realistic PFS–OS surrogacy levels for PM.

For the power models, the outcome was a plot of misclassification rate against the resulting proportion of trials that would generate a positive result, with 0% misclassification resulting in 80% positive trials based on the intended 80% power being delivered. For the endpoint precision models, the outcome was a plot of misclassification rate against the proportion of observed endpoint CIs that actually contained the result for each trial. With 0% misclassification, 95% of the modelled trials contained the true effect size, as intended.

For all models, misclassification (incorrect classification relative to true response) was used rather than discordance (disagreement between two reviewers) since the former directly corresponds to clinical practice and clinical trials where only one report is acquired (whether this is a centralised or local opinion). The additional assumptions needed to model discordance rate directly added considerable complexity but no additional clinical value. For comparison between the primary analysis and the *in silico* modelling, the discordance rate observed in the primary analysis was halved to define a conservative lower bound of the misclassification rate observed in the retrospective study cohort, since by definition, at least one of two discordant reports must be wrong, if there is only true response state.

#### Root Cause Analysis in Discordant cases

Discordant cases were classified into those spanning adjacent response groups (PR v SD, SD v PD) and those spanning PR v PD. Root cause analysis involved allocation of each discordant case to one of the following *a priori* groups: subjective difference (contrasting interpretation with no definitive identifiable cause), human error (incorrect application of mRECIST criteria, missed imaging finding(s), incorrect CT imaging reviewed, incorrect data recording) and variances related to mRECIST measurements (defined as <10% difference in summed values following correct application).

#### Interaction between Discordance Rate and Tumour Volume

A subset of cases underwent volumetric tumour annotation for AI training, as recently reported^10^. This facilitated comparison between baseline volume and volumetric response relative to discordance rate and mRECIST measurements.

#### Statistics

A sample size calculation was not performed given the exploratory nature of the study. Data are described by simple proportions or by mean (SD)/median (IQR) based on distribution. Unpaired t-tests or Mann-Whitney U-tests were employed for subsequent comparisons. Fisher’s exact test was used to compare discordance rates between sites and to assess interactions with tumour volume. Cohen’s kappa was used to describe mRECIST inter-observer agreement. Modelling and waterfall plots were generated in R, v4.5.2 and Python, v 3.10.12. Other calculations were performed using SPSS v 28.0.

## RESULTS

### Study Population & Image Acquisition

172 cases were included. Mean age was 68 (7) years. 142/172 (83%) were male. Histological subtype was epithelioid in 131 (76%), sarcomatoid in 21 (13%), biphasic in 13 (8%) and unrecorded in 7 (4%). Chemotherapy intent was palliative in 131 patients (76%) and neoadjuvant (pre-surgery) in 41 (24%). 132/172 (77%) had venous-phase contrast-enhanced CT, 33/172 (19%) had arterial-phase contrast CT and 7/172 (4%) had non-contrast imaging.

### Primary Objective: Discordance Rate

Response classifications by reporters 1 and 2 are summarised in Figure 1. This demonstrates that mRECIST classifications were discordant in 60/172 cases, meaning the discordance rate was 35%. Inter-observer agreement was only moderate (k=0.456 (0.346-0.566)) and was not different between sites (p=0.198). Discordance rate did not differ by site (Glasgow: 37/90 (41.1%), Leicester: 14/49 (28.6%), Wythenshawe: 9/33 (27.3%), p=0.295).

**Figure 1.**
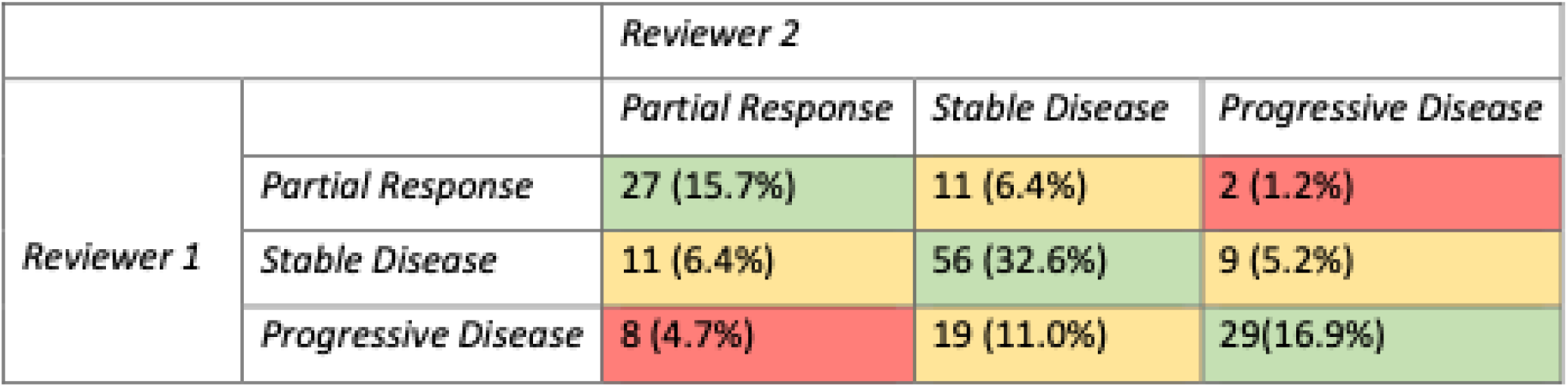
Confusion matrix summarising response classification in 172 patients with Pleural Mesothelioma treated with chemotherapy based on independent review by two expert radiologists using modified RECIST version 1.1. Classifications were concordant in 112/172 (65%), highlighted in green. Response classification was discordant in 60/172 (35%), including 50 classifications that spanned adjacent response groups (SD v PR, SD v PD), highlighted in amber, and 10 classifications that spanned PD v PR, highlighted in red.

### Secondary Objectives

#### In Silico Modelling of Impact on Clinical Trials

Increasing misclassification rate was associated with a progressive decline in the power (see Figures 2A-D) and endpoint coverage (see Figures 3A-D) delivered in clinical trials across all four endpoints. At a misclassification rate of 17%, which approximates to a conservative lower bound estimate of this ‘single reporter’ value when 35% of cases are discordantly reported by two reporters, statistical power dropped from 80% to 55% for ORR, 53% for DCR, 65% for PFS and 66% for OS (see Figure 2A-D, respectively). At 17% misclassification we observed corresponding reductions in endpoint coverage to 88% for ORR, 89% for DCR, 92% for PFS and 92% for OS, (see Figure 3A-D, respectively).

**Figure 2.**
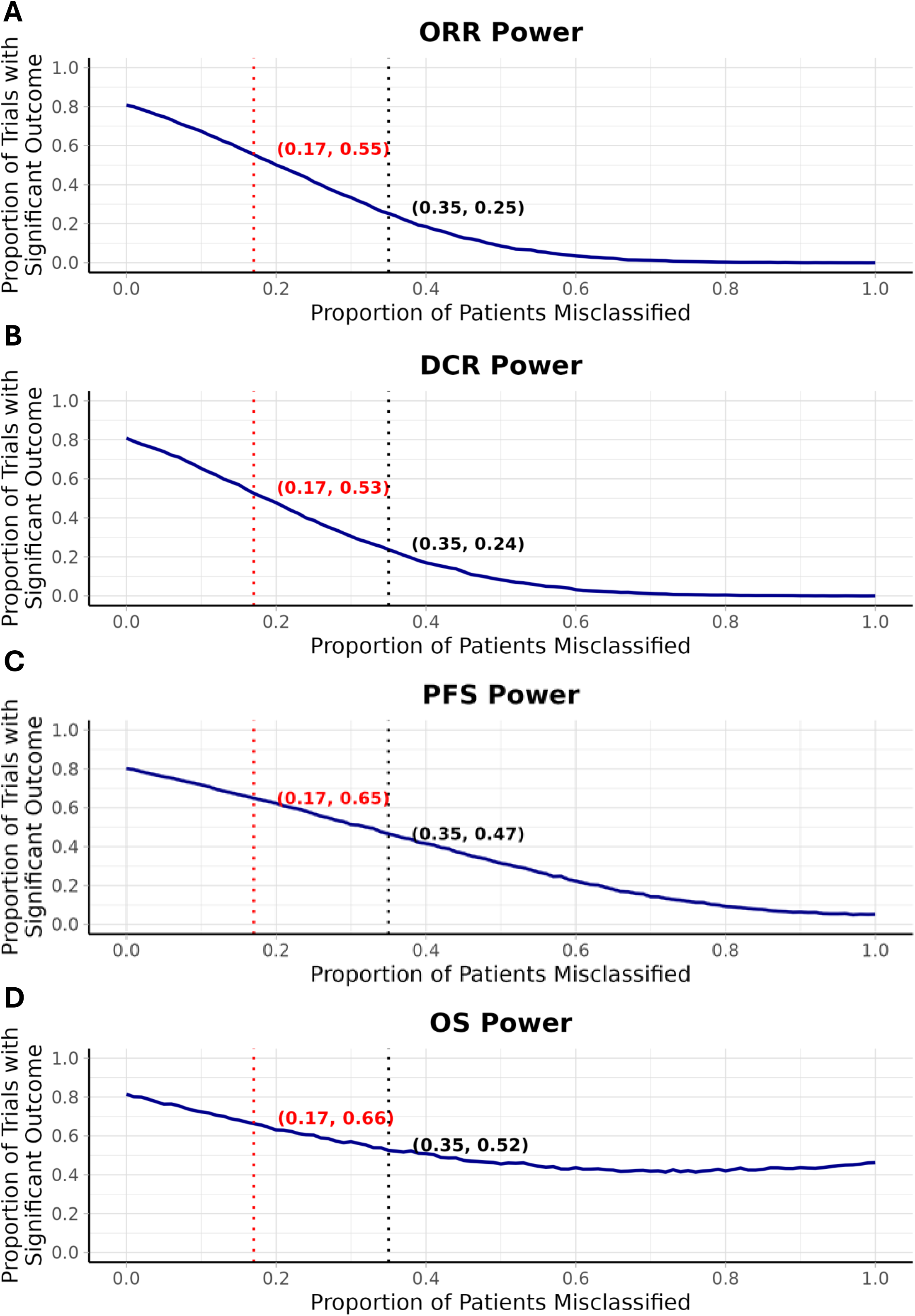
Effect of patient misclassification on the statistical power for trials investigating ORR (A) and DCR (B) when powering for true absolute differences of 20%, and OS (C) when powering for a true effective hazard ratio of 0.7. 80% power and a significance level of 0.05 are targeted and misclassification rate is varied from 0% to 100%. 10,000 simulated trials were run per misclassification proportion. Across all endpoints, power declines monotonically with increasing misclassification.

**Figure 3.**
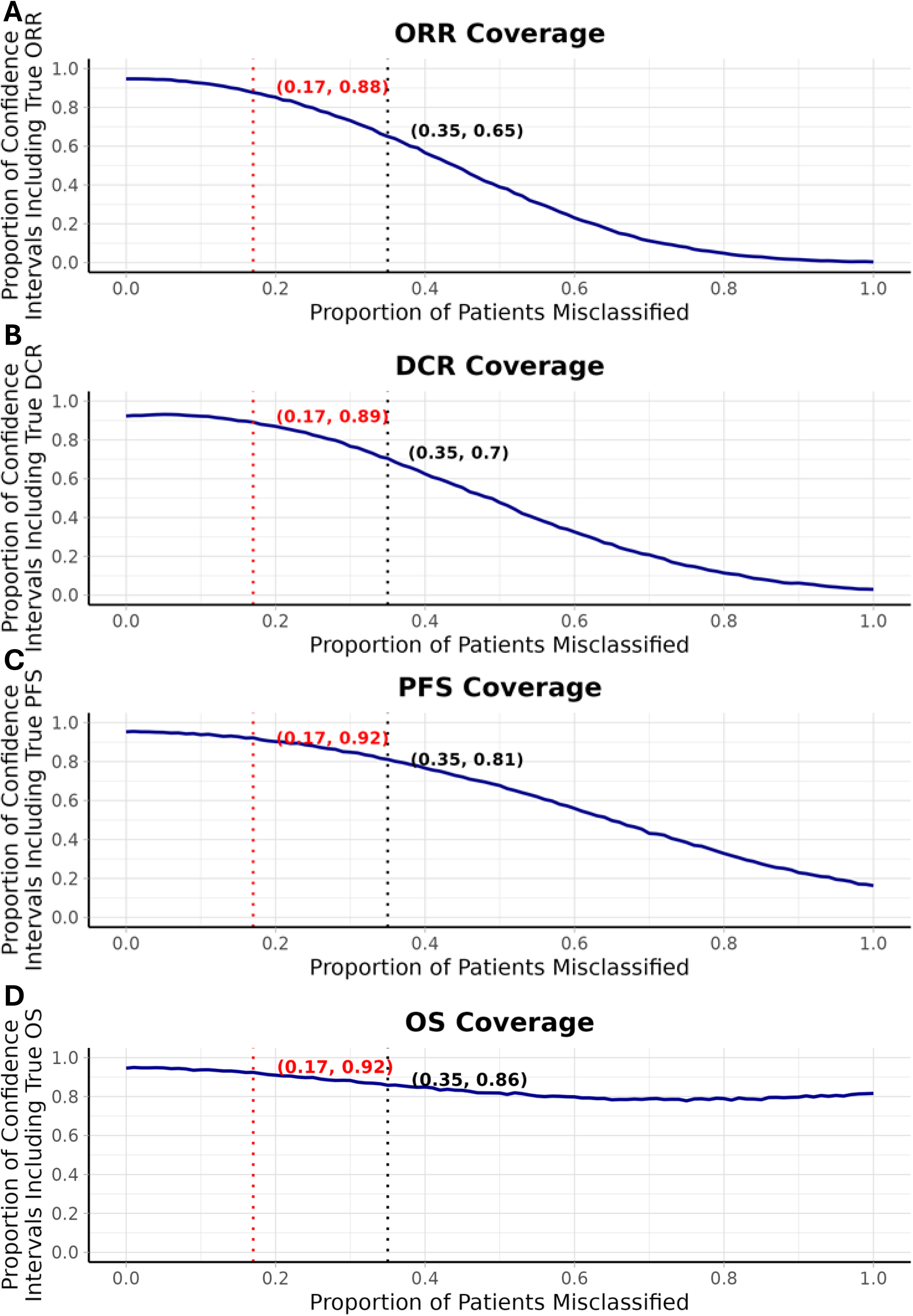
Effect of patient misclassification on the confidence interval precision for trials investigating ORR (A) and DCR (B) when powering for true absolute differences of 20%, and OS (C) when powering for a true effective hazard ratio of 0.7. 80% power and a significance level of 0.05 are targeted, and misclassification rate is varied from 0% to 100%. 10,000 simulated trials were run per misclassification proportion. Across all endpoints, confidence interval precision declines monotonically with increasing misclassification.

#### Root Cause Analysis of Reasons for Discordant Classification

50 of the 60 discordant classifications (83%) spanned adjacent groups (PR v SD, SD v PD). The 10/60 (17%) that spanned PR v PD all reflected human errors, including review of the wrong imaging (7/10), incorrect application of mRECIST (2/10) and a missed finding (1/10). In the remaining 50 discordant cases that spanned one response group, 24/50 (48%) reflected subjective differences, 14/50 (28%) reflected variations in calliper placement during mRECIST measurement and 12/50 reflected human errors (4 (8%) incorrect application of mRECIST, 4 (8%) missed finding, 2 (4%) recording errors, 2 (4%) wrong imaging reviewed).

#### Interaction between Discordance Rate and Tumour Volume

70/172 cases (41%) had baseline volumetry available. 24/172 (14%) had baseline and response CT volumetry available.

##### Baseline Tumour Volume

The median baseline tumour volume was 360 cm^3^. As illustrated in Figure 4, discordance rate was not associated with baseline volume. When dichotomised around the median value, the odds ratio (OR) of discordance was not higher in cases with lower volume (1.07 (95% CI 0.41-2.7). Results were similar in a sensitivity analysis in which 20 cases with extra-pleural disease (nodal or distant metastases) or simple human errors were excluded to remove confounding sources of discordance (OR 1.0 (95% CI 0.31-3.28)).

**Figure 4.**
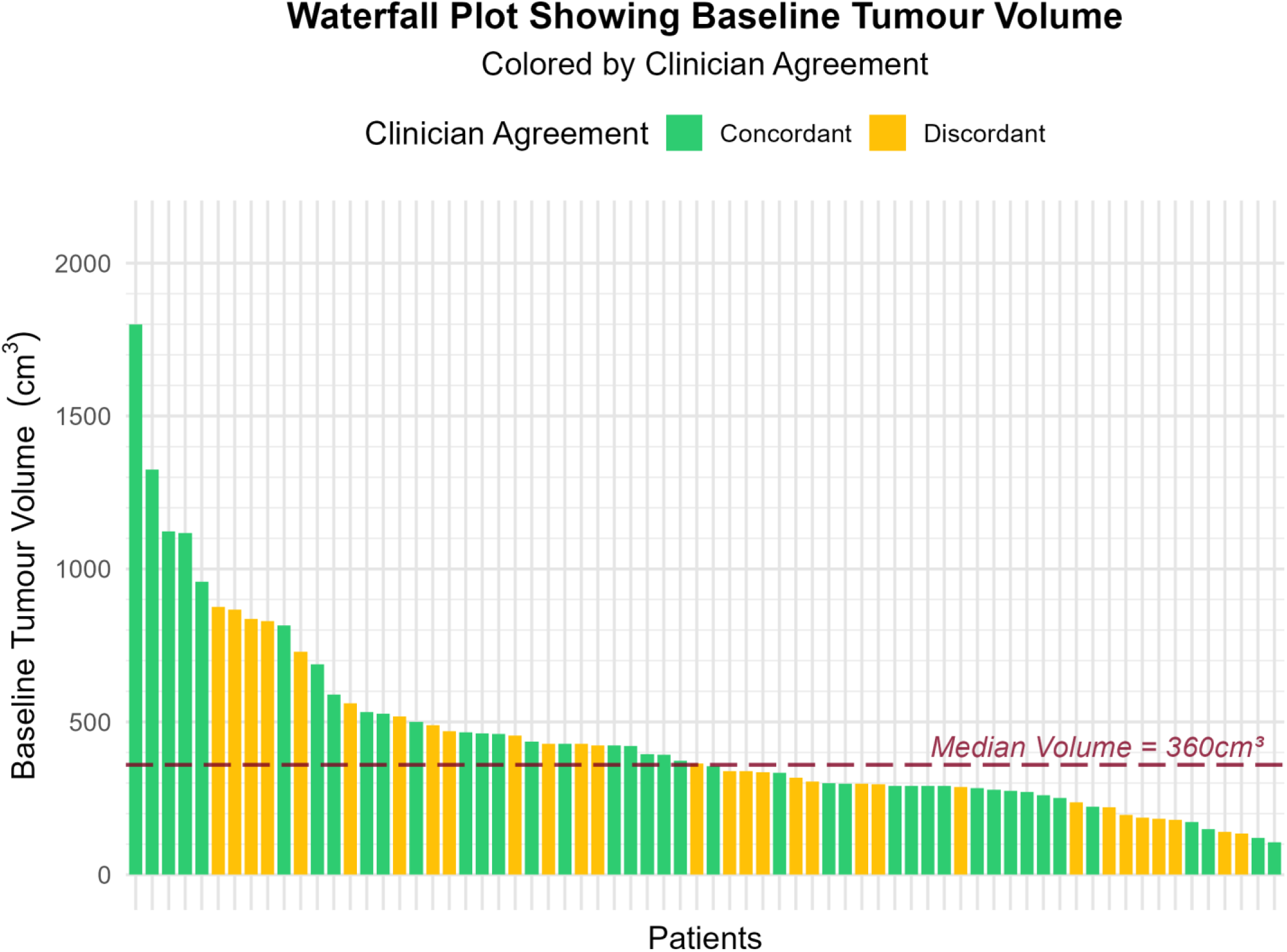
Histogram showing the interaction between baseline tumour volume and discordant mRECIST reporting in 70/172 patients with baseline volumetry available. The red line corresponds to the median baseline tumour volume (360 cm^3^). Discordance rate was not different in cases with baseline volumes above and below 360 cm^3^ (both 8/50, 16%, p=1.0).

##### Volumetric Response

Pleural tumour volume fell (Δ volume <0 cm^3^) during chemotherapy in 15/24 (62.5%) patients with available baseline and response volumes. As shown in Figure 5, there was poor agreement between volumetric response and mRECIST classification. Similar findings were observed in a sensitivity analysis excluding 4 cases with extra-pleural disease and 2 cases in which misclassification reflected image selection/processing errors. Discordance rate was not different in cases with volumetric response v progression (5/15 (33%) v 2/9 (22%) respectively (p=0.669)). A similar result was observed when cases with extra-pleural disease or image selection/processing errors were excluded (4/10 (40%) v 1/8 (13%), p=0.314).

**Figure 5.**
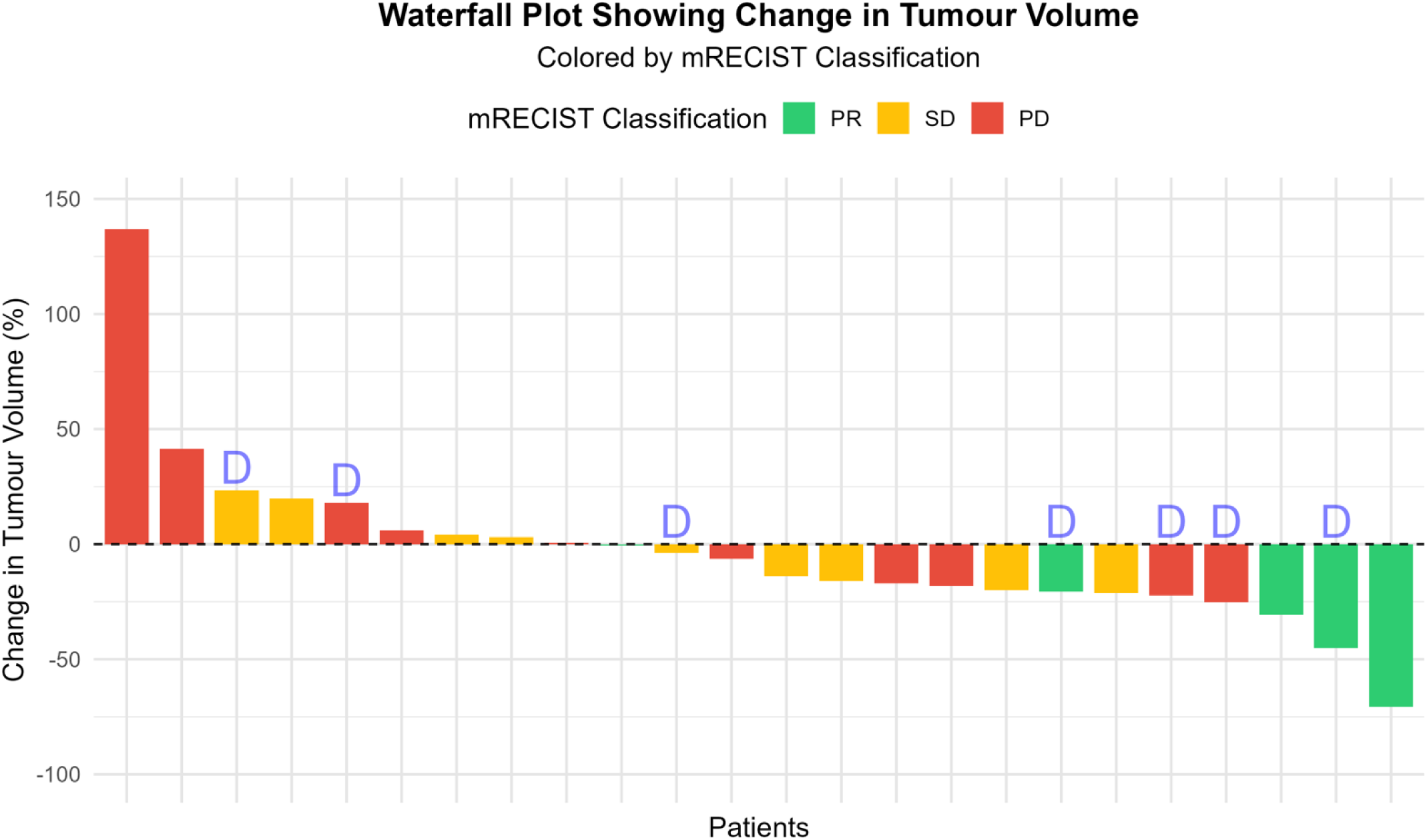
Waterfall plot showing the interaction between volumetric change (Δ), mRECIST response and instances of discordant mRECIST reporting (Bars labelled ‘D’) in 18/172 study cases in which manual volumetry was performed in both baseline and response imaging. Discordant cases that reflected human error (n=2) and cases with nodal or extra-pleural disease measurements (n=4) have been excluded. Red bars correspond to progressive disease, yellow bars correspond to stable disease, green bars to partial response. Higher discordance rate was observed when tumour volume reduced (4/10 (40%), compared with 1/8 (13%) in cases with positive Δ volume values (p=0.444). Cases with volume reduction were also more at risk of discordant reporting in our sample (OR 4.67, 95% CI 0.4-54.0).

## DISCUSSION

In this retrospective, multicentre study of 172 chemotherapy-treated PM patients, we found that expert radiologists disagreed regarding mRECIST response classification in 35% of cases. The level of agreement between radiologists was therefore only moderate (k=0.456 (0.346-0.566)). In a root cause analysis, most discordances (50/60, 83%) reflected subtle image interpretation or measurement differences intrinsic to the application of mRECIST. The remaining minority reflected human error, including incorrect selection of the appropriate CT from a larger series. *In silico* modelling of the impact of erroneous response classification demonstrated sizeable reductions in the intended statistical power and endpoint coverage of clinical trials as misclassification rate increased. The modelling experiments we conducted used ‘misclassification rate’ as a single reporter equivalent of discordance rate, which was used in the primary analysis. Importantly, calculation of discordance rate requires two reporters, at least one of whom must, by definition, be incorrect for discordance to occur. In our *in silico* models, the statistical power of a hypothetical two-arm trial of an active PM therapy, dropped from intended levels of 80% to 53-66%. At the same misclassification rate, endpoint coverage narrowed from intended 95% confidence intervals to coverage of only 88-92% of the true treatment effect.

Discordance rate between expert mRECIST reporters has not previously been reported in the literature. However, our observations are consistent with previous studies that reported differences in individual mRECIST measurements of up to 30% between radiologists^4^ . Our data suggest that these measurement variances frequently translate into different final mRECIST classifications. This is perhaps unsurprising since a measurement variance of 30% exceeds the established mRECIST thresholds for PD and SD (≥20% and ≤30%, respectively).

The unique growth pattern of PM presents considerable challenges for accurate and consistent tumour measurement on different scans by human readers^11,12^ . As previously summarised, these challenges persist despite improvements incorporated in the current version 1.1 of mRECIST^13^ . PM is typically disseminated across a large pleural surface area, often with minimal cross-sectional thickness. This makes acquisition of transverse unidimensional measurements, as required by mRECIST, exceptionally challenging, with subtle differences in calliper angulation and timing resulting in different values^14,15^ . In a previous PM study, lesion size varied by an average of 15.1% across 170 pleural locations when measured by a single expert radiologist^15^ . Similar systematic error of measurement is reported in pulmonary nodule assessment^16^ , where the inherent contrast between the target lung nodule and surrounding lung parenchyma is considerably higher. Some of the modifications made in mRECIST v.1.1, including a minimum measurement of 2mm (instead of the standard 5mm), may also have increased the probability and extent of inter-observer variation and consequent divergence in response classification.

Heterogeneous tumour response may also complicate response assessment since failure to perfectly replicate calliper positions on follow-up imaging may produce accurate but divergent growth patterns. More holistic, volumetric assessment is an obvious solution to mitigate this. In a cohort of 116 PM patients undergoing extended pleurectomy and decortication, resected tumour volume was a significant predictor of survival^17^ . In a small retrospective study (n=30), Frauenfelder et al previously demonstrated that volumetric response criteria reclassified some cases from mRECIST-PR and -SD to SD and PD, respectively. In this study, volumetric response was also associated with higher interobserver agreement than mRECIST (k = 0.9 v 0.33)^18^ . Unfortunately, a subsequent large, multi-centre volumetry study, demonstrated that, like mRECIST, human-generated tumour volumetry was also confounded by inter-observer variation, with a mean volume difference between radiologists of 47.9ml (+/- 178.2) cm^3^. In this study, Gill et al reported root causes for inter-observer disagreement that were similar to those reported here, including data entry error, user error, and perception error including those related to poor distinction between tumour and adjacent structures on CT^11^ . In our opinion, successful deployment of recently reported AI solutions for automated tumour segmentation^10,19^ offers the greatest potential to mitigate these errors in human generated manual volumetry. Use of a dedicated radiology reporting template, ideally combined with AI, may also address the user errors and inconsistences reported here, and by Gill et al. A reporting template for PM pathology has been published^20^ and in non-PM radiology, Jorg et al, recently reported enhanced satisfaction among both radiologists and requesting physicians using this approach^21^ .

### Implications for Clinical Trials and Routine Practice

The *in silico* modelling outcomes reported here suggest that, if similar vulnerabilities exist in real reporting pipelines, which seems likely given previously reported evidence of inferior radiology reporting performance from non-specialists in thoracic oncology^22^ , clinical trials are operating with significantly reduced statistical power and endpoint coverage than intended. In a rare cancer like PM, this argues strongly for adoption of endpoints less vulnerable to human error, including AI-assisted PFS and OS, although use of the latter is challenging when multiple lines of therapy may confound post-protocol effects.

In current clinical practice, combination immunotherapy using ipilimumab and nivolumab is the licensed standard of care for patients with later stages of diseases (defined as ‘unresectable’ disease). However, real-world studies have failed to replicate the licensing trial (Check Mate743^2^ ), with a median OS of only 13.7 months^23–25^ compared to 18 months in Check Mate 743. In the real world, Ipi-Nivo also appears relatively poorly tolerated, with any grade adverse effects experience by >60% patients^24^ and 1-in-4 patients stopping treatment due to toxicity^24,25^ . Chemotherapy has also proven disappointing in a real-world setting, with <25% patients responding^26,27^ . This apparently poor calibration between trial results and real-world outcomes may in part be explained by inconsistent radiological response assessment in either the clinical trial, the real world or both.

### Study Limitations

It is important to acknowledge some key limitations of the current study including its retrospective design and relatively small size. Nevertheless, to our knowledge, the current report represents the first multicentre systematic examination of real-world performance of mRECIST. The themes used to report sources of error in the root cause analysis are intrinsically subjective. In addition, the requirement for each reporting radiologist to first identify and retrieve the correct scan for Glasgow cases only from the national PACS system may also have biased these cases towards more incorrect study selection. Baseline and response scans from other centres were simply presented to each radiologist on an encrypted hard drive. However, this highlights this important source of error, which most closely replicates how radiologists perform reporting in clinical practice. The number of cases with available volumetry data was limited, which reduces the statistical power available to assess any true interaction with discordance rate.

Due to the patient-level assumptions inherent in the coverage and power modelling for OS, these simulations which assume no long-lasting treatment effect are less representative of immunotherapies because of their durable survival benefits. However, both power and coverage would still be expected to decrease under these treatments. Additionally, we acknowledge that the probability of misclassification is unlikely to be the same for each of the mRECIST classes despite our assumption; notably, in this study we observed that patients PD-SD disagreements were slightly more common than PR/SD disagreements (28/60 vs. 22/60), and both were more frequent than PD/PR disagreements (10/60).

## CONCLUSION

This retrospective multicentre study demonstrates considerable discordance between expert PM radiologists in the assessment of treatment response. Different response classification was observed in 35% of cases and inter-observer agreement was only moderate (k=0.456 (0.346-0.566)). *In silico* modelling demonstrated the quantitative impact of erroneous response assessment for the first time, including considerable reductions in theoretical statistical power and endpoint coverage. These findings highlight the urgent need to embed better endpoints for clinical trials and routine clinical care, including AI-assisted measurements of tumour growth.

## Data Availability

Data are available on request to the corresponding author or via PREDICT-Meso.

